# Perioperative outcomes in patients with myalgic encephalomyelitis/chronic fatigue syndrome undergoing general anesthesia: a retrospective matched-pair study

**DOI:** 10.64898/2026.04.06.26348924

**Authors:** Felix M. Steinkirchner, Christina K. Kaufmann, Richard F. Kraus, Maximilian Käss, Elisabeth Schieffer, Bernhard M. Graf, Christoph Lassen, Viktoria Kimmerling, Alexander Dejaco

**Affiliations:** Department of Anesthesiology, ME/CFS Research Group, University Hospital Regensburg, Regensburg, Germany; Department of Cardiology, Angiology and Intensive Care, University Hospital Gießen and Marburg, Marburg, Germany

**Author notes:** Corresponding author: E-Mail address, Postal Address: Dr. F. Steinkirchner, Department of Anesthesiology, University Hospital Regensburg, Franz-Josef-Strauß-Allee 11, 93053 Regensburg, Germany.

**Keywords:** ME/CFS, Myalgic Encephalomyelitis, Chronic Fatigue Syndrome, General Anesthesia, Intraoperative Hypotension, Postoperative Pain, Dysautonomia

## Abstract

**Background:** Myalgic encephalomyelitis/chronic fatigue syndrome (ME/CFS) is a chronic multisystem disease characterized by profound fatigue, post-exertional malaise, cognitive impairment, and autonomic dysfunction. Its pathophysiology is incompletely understood and likely involves complex interactions between immune, autonomic, and metabolic dysregulation. Despite features with potential relevance for anesthesia and perioperative care, evidence to guide anesthetic management in individuals with ME/CFS remains limited. We therefore performed a retrospective matched-pair analysis to generate clinical data on perioperative responses and identify areas for future research.

**Methods:** We conducted a retrospective matched-pair analysis at a single tertiary center. All patients with ME/CFS undergoing general anesthesia from 2015 to 2026 were identified using ICD-10-GM codes (G93.3 and U09.9) with additional manual verification and matched 1:1 to controls for comparison. Patients with confounding diagnoses or American Society of Anesthesiologists physical status above III were excluded. The analysis focused on intraoperative hemodynamic parameters, including baseline, post-induction, median, and lowest recorded systolic blood pressure and heart rate, as well as early postoperative outcomes in the post-anesthesia care unit (PACU), including maximum pain scores and requirement for rescue analgesia.

**Results:** Out of 189 individuals identified through ICD-10 codes, 15 matched pairs were included after application of exclusion criteria. ME/CFS patients exhibited lower lowest recorded intraoperative systolic blood pressure (90.0 [82.5-95.0] mmHg in ME/CFS vs 100.0 [90.0-110.0] mmHg in controls, p = 0.044) as well as lower lowest heart rate (50 [40.0-57.5] bpm in ME/CFS vs 60 [50.0-65.0] bpm in controls, p = 0.012). Vasopressor use and fluid administration did not differ, and no episodes of severe hypotension or perioperative adverse events were observed. Postoperative pain was higher in ME/CFS, with higher maximum pain scores (NRS 5.0 [4.0-6.0] in ME/CFS vs 1.0 [0.0-4.0] in controls, p = 0.008) and more frequent opioid rescue analgesia (80% in ME/CFS vs 33% in controls, p = 0.039). Postoperative nausea or vomiting, oxygen supplementation, and PACU length of stay were similar between groups.

**Conclusions:** General anesthesia appears hemodynamically well tolerated in individuals with ME/CFS. In contrast, postoperative pain burden is increased and may require tailored analgesic strategies. Post-exertional malaise, a key disease feature with potentially delayed onset and significant impact, was not captured in this study and remains an important target for future research. These hypothesis-generating findings highlight the need for prospective studies to optimize perioperative management and evaluate patient-relevant outcomes in ME/CFS.

## 1. Introduction

Myalgic encephalomyelitis/chronic fatigue syndrome (ME/CFS) is a chronic multisystem disease characterized by post-exertional malaise (PEM), debilitating fatigue, cognitive dysfunction, and dysautonomia [1], affecting an estimated 0.68% of the population [2]. The pathophysiology remains incompletely understood and likely involves complex interactions between autonomic, immune, and metabolic dysfunction [3–5].

Several features of ME/CFS have direct relevance for perioperative care. Autonomic dysfunction with altered cardiovascular regulation is common and frequently manifests as orthostatic intolerance (OI) or postural orthostatic tachycardia syndrome (POTS), characterized by inappropriate vascular tone and exaggerated tachycardic responses [6]. A relevant subset of patients experiences OI or POTS, which may lead to suboptimal hemodynamic responses under anesthesia [7,8]. Anesthetic agents inherently induce vasodilation and myocardial depression, effects that may be amplified by pre-existing autonomic dysregulation [9,10]. This may predispose to hypotension, impaired tolerance to hypovolemia, or abnormal cardiovascular responses to surgical positioning and conditions that reduce venous return, such as ventilation with positive end-expiratory pressure, steep anti-Trendelenburg or pneumoperitoneum during laparoscopic procedures [11,12]. Neuropathy and heightened nociceptive processing, including mechanisms such as central sensitization, may contribute to increased perioperative pain perception and greater analgesic requirements, with a subsequent higher risk of related adverse effects [13–15]. Gastrointestinal dysmotility and delayed gastric emptying have been reported in subsets of patients, potentially affecting perioperative medication handling and aspiration risk [16,17]. Immune dysregulation and abnormal mast-cell activity have been proposed as contributors to symptom generation and could theoretically influence perioperative inflammatory responses or hypersensitivity-like reactions [4,18–20]. Dysautonomia and neuromuscular fatigue could theoretically affect airway reflexes and recovery from anesthesia, potentially manifesting as prolonged emergence, delayed extubation, or increased risk of respiratory complications. Finally, PEM - a defining feature of ME/CFS - raises the possibility of impaired postoperative recovery and prolonged functional decline after surgical stress [21,22].

Available evidence of the perioperative impact of ME/CFS is limited and consists largely of small case series and expert opinion [21,23–27]. In the only published clinical case series to date, Fisher and Rose reported perioperative outcomes in 27 individuals with a history or perceived risk of adverse reactions after anesthesia in the context of chronic fatigue syndrome or idiopathic environmental intolerance, describing postoperative symptom exacerbations but no major intraoperative complications [27]. However, this cohort is unlikely to adequately represent the baseline ME/CFS population and did not include a control group.

Consequently, robust clinical evidence to guide perioperative management or anesthetic techniques in individuals with ME/CFS remains lacking. To date, no study has systematically examined intraoperative hemodynamic stability or postoperative pain in a matched cohort of patients with ME/CFS.

Despite this, ME/CFS remains markedly under-represented in the anesthesia literature, and perioperative care is largely delivered as “care as usual” rather than being informed by disease-specific evidence. This represents a substantial unmet clinical need, as the absence of robust data limits the ability to identify perioperative factors that may exacerbate symptoms and hinders the development of tailored anesthetic strategies. Generating clinical data in this area is therefore essential to characterize perioperative responses in ME/CFS and to inform future prospective studies evaluating modifiable anesthesia-related factors, including choice of anesthetic technique.

We therefore conducted a retrospective matched-pair analysis to characterize perioperative responses, including intraoperative hemodynamic parameters and postoperative pain, in patients with ME/CFS compared with matched controls undergoing general anesthesia.

## 2. Methods

### 2.1. Study Design and Patient Identification

We conducted a retrospective single-center analysis at the University Hospital Regensburg covering the period from January 2015 to January 2026. All patients with a documented diagnosis of ME/CFS (International Classification of Diseases 10th Revision, German Modification (ICD-10-GM) code G93.3) or post-COVID syndrome (ICD-10-GM code U09.9) and a recorded surgical procedure were identified through the institutional clinical information system (CIS).

For patients coded with G93.3, the diagnosis was accepted as established without further verification. For patients coded with U09.9, medical records, including discharge letters, were manually reviewed, as this code encompasses a heterogeneous post-COVID population in which ME/CFS is not uniformly present. This step was necessary because a proportion of patients with ME/CFS in the institutional CIS were coded exclusively as U09.9 without an accompanying G93.3-code, while the majority of U09.9-coded patients did not fulfill criteria for ME/CFS. Consequently, all included cases in the subsequent analyses had a physician-documented diagnosis of ME/CFS at the time of anesthesia, either formally coded as G93.3 or identified through manual review of medical records in patients coded exclusively as U09.9. Formal diagnostic criteria such as the Canadian Consensus Criteria [22] were not applied.

### 2.2. Exclusion Criteria

Cases were excluded if they had not undergone general anesthesia for their procedure, if confounding diagnoses were present (malignant, neurodegenerative, or decompensated illness, operationalized as an American Society of Anesthesiology (ASA) physical status score > III), or if the ME/CFS diagnosis had only been established after the surgical procedure. If multiple procedures were recorded for a patient, only the most recent procedure was included, as ME/CFS diagnosis could not in every case be assumed to have been present at the time of earlier procedures.

### 2.3. Matching

Each eligible case with ME/CFS was matched 1:1 to a control subject without ME/CFS using two formal criteria: sex and surgical procedure (same or clinically comparable procedure). Additional variables were not used as formal matching criteria to avoid over-restriction of eligible pairs. Instead, relevant baseline and procedure related characteristics, including age, body mass index (BMI), anesthetic technique, and year of surgery, were assessed post hoc to evaluate comparability between groups.

### 2.4. Data Extraction

Data were extracted from hand-written anesthetic, post-anesthesia care unit (PACU) records and discharge letters. Intraoperative vital signs were routinely recorded at five-minute intervals. Extracted parameters included intraoperative fluid use, vasopressor use (maximum noradrenaline infusion rate during continuous infusion), total administered anesthetic and opioid dose and averaged opioid dose per time, duration of anesthesia, time from end of surgery to extubation, and hemodynamic trajectories.

### 2.5. Endpoints

In keeping with the exploratory nature of this study, no single primary endpoint was designated; instead, two pre-specified domains were defined a priori as the focus of the analysis: intraoperative hemodynamic stability, assessed through post-induction, median intraoperative, nadir, and lowest sustained values for both systolic blood pressure and heart rate, as well as postoperative pain intensity, defined as the highest numerical rating scale (NRS) pain score recorded in the PACU. Baseline blood pressure and heart rate were defined as the last recorded values prior to anesthetic induction. The intraoperative nadir was defined as the single lowest value recorded at any time point; the lowest sustained value as the lowest value confirmed across consecutive measurements spanning at least 15 minutes.

Secondary outcomes included vasopressor requirements, intraoperative opioid consumption, time to extubation, PACU stay duration, opioid rescue analgesia, defined as any opioid administration in the PACU (piritramide, oxycodone, or hydromorphone), non-opioid rescue analgesia (metamizole, ibuprofen, parecoxib, paracetamol), postoperative nausea and vomiting (PONV), supplemental oxygen requirement, vital signs in the PACU at admission and discharge, and postoperative hospital length of stay.

### 2.6. Statistical Analysis

Statistical analysis was performed in R (version 4.5.1; R Foundation for Statistical Computing, Vienna, Austria). Matched pairs were compared using the Wilcoxon signed-rank test for continuous variables and McNemar’s test for binary outcomes. A p-value < 0.05 was considered statistically significant. No formal sample size calculation was performed; the study included all eligible patients identified during the study period. This approach was chosen because the study aimed to describe the complete available experience at a single center over an eleven-year period, and no prior data existed on which to base a meaningful power calculation. Variables with missing values are indicated in the respective tables; no imputation was performed. Continuous variables were analyzed on their original scale without categorization. No subgroup analyses, interaction tests, or formal outlier exclusion were performed given the small sample size; individual data points are displayed in all figures. No sensitivity analyses were performed.

### 2.7. Potential sources of bias

Several potential sources of bias inherent to this study design should be acknowledged. First, ICD-10-based case identification may result in both false inclusions (miscoded patients) and false exclusions (undiagnosed or uncoded ME/CFS); residual confounding from this source cannot be fully excluded. Second, severity selection bias is inherent to a surgical cohort: patients with the most severe ME/CFS are less likely to undergo elective procedures and are therefore systematically under-represented. Third, the five-minute interval manual hemodynamic documentation introduces measurement imprecision, particularly for transient hemodynamic events.

## 3. Results

### 3.1. Study population and perioperative characteristics

Extraction of cases based on ICD-10 codes yielded 75 and 114 patients who underwent a surgical procedure with a diagnosis of G93.3 or U09.9, respectively. From those with a G93.3 diagnosis, 20 were excluded because no general anesthesia was performed, and 42 were excluded due to malignancy, terminal illness, or ASA status > III. Of the remaining 13 cases, 2 were excluded because the diagnosis of ME/CFS had only been established after the surgical procedure and was therefore not present at the time of anesthesia, leaving 11 eligible cases in the G93.3 cohort. Out of patients with a U09.9 diagnosis, 92 were excluded as ME/CFS was not confirmed in their medical records, 7 because no general anesthesia was performed, 10 due to malignancy, terminal illness, or ASA status above III, and 1 because the ME/CFS diagnosis had only been established after the surgical procedure. This left 4 eligible patients in the U09.9 cohort. This resulted in 15 eligible ME/CFS cases overall, which were matched with 15 controls (Fig. 1). All subsequent analyses refer to this cohort.

**Fig. 1:**
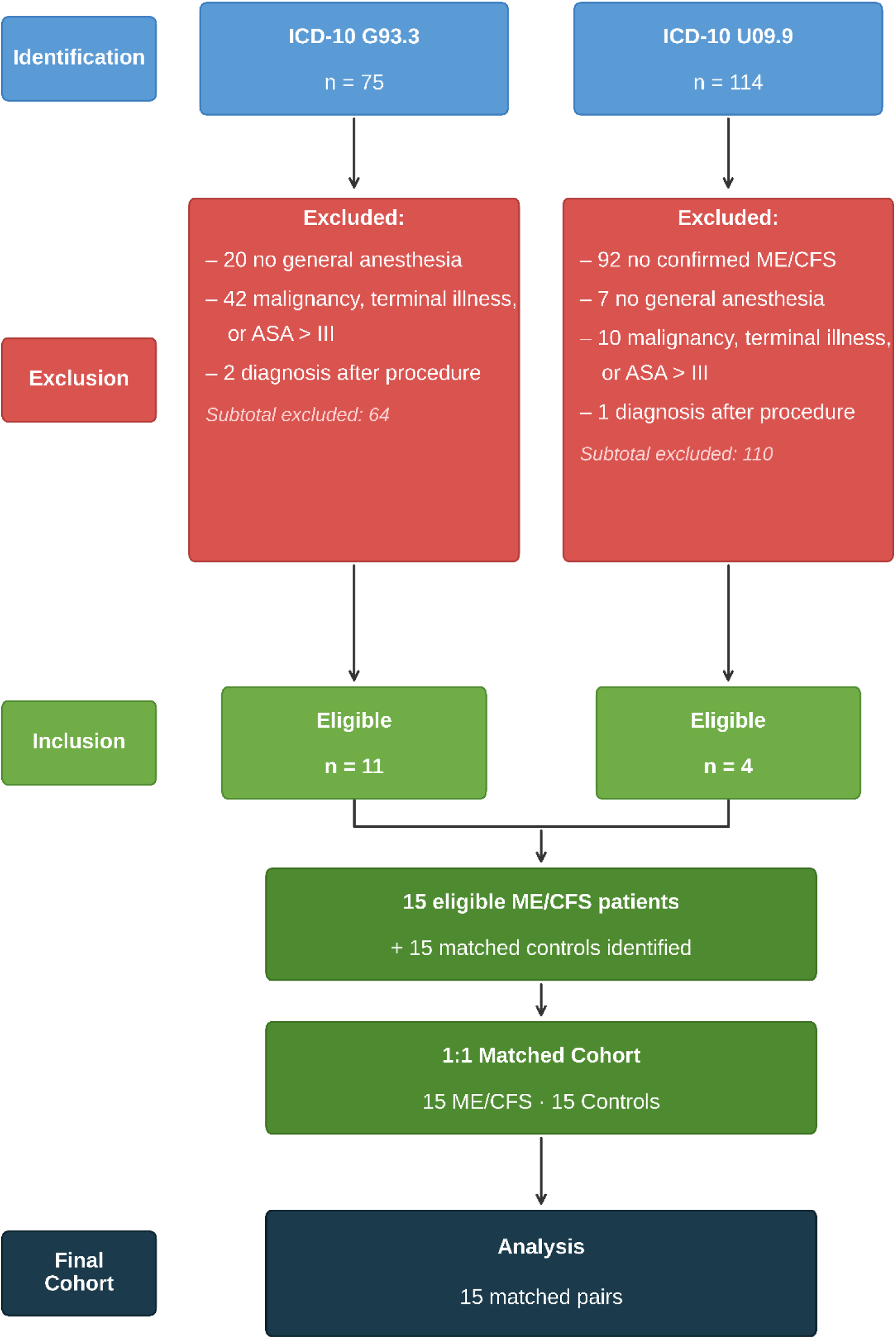
Flow diagram of participant identification and selection. ICD-10 = International Classification of Diseases, 10th Revision, German Modification; ASA = American Society of Anesthesiologists; ME/CFS = myalgic encephalomyelitis/chronic fatigue syndrome. Controls were matched 1:1 by sex and surgical procedure type. Where a patient had undergone multiple procedures, only the most recent was included. Patients may fulfill more than one exclusion criterion; only the first applicable criterion was counted.

No eligible cases were found between 2019 and 2021, coinciding with the COVID-19 pandemic and the associated reduction in elective surgical activity. Case numbers increased markedly from 2022 onwards, paralleling the rising clinical awareness of ME/CFS in the context of post-COVID.

Baseline characteristics were comparable between groups (Table 1). No significant differences were observed for any baseline characteristic (all p ≥ 0.196), except for the year of the procedure (p = 0.003). The cases in the ME/CFS cohort were unevenly distributed across the study period, however, matched cohorts were well aligned (see Supplemental Figure S1). Out of 15 procedures, 11 (73.3%) took place within one year of each other, with a median offset of 8.3 months (IQR 3.1-15.8). Surgical procedures were heterogeneous overall, but also well aligned between matched cohorts (Supplemental Table S1).

**Table 1.** Baseline characteristics.

| Variable | ME/CFS (n=15) | Control (n=15) | p-value |
| --- | --- | --- | --- |
| **Demographics** |  |  |  |
| Age (years) | 47.7 (41.8–60.3) | 49.5 (37.8–61.7) | 0.629 |
| Female sex, n (%) | 11/15 (73%) | 11/15 (73%) | — |
| BMI (kg/m<sup>2</sup>) | 29.7 (23.9–34.5) | 26.9 (25.0–31.0) | 0.887 |
| Weight (kg) | 86.0 (67.0–105.0) | 80.0 (68.5–90.5) | 0.267 |
| Height (cm) | 174.0 (165.0–178.0) | 168.0 (161.5–175.0) | 0.196 |
| **Clinical status** |  |  |  |
| ASA status (I/II/III) | 0/8/7 | 1/9/5 | — |
| History of PONV, n (%) | 4/15 (27%) | 2/15 (13%) | 0.687 |
| Preoperative anxiolysis, n (%) | 2/15 (13%) | 3/15 (20%) | 1.000 |
| **Procedure** |  |  |  |
| Additional regional anesthesia, n (%) | 1/15 (7%) | 1/15 (7%) | — |
| Anesthesia duration (min) | 115.0 (95.0–130.0) | 100.0 (67.5–160.0) | 0.459 |
| Year of procedure | 2024 (2020–2024) | 2025 (2022–2025) | 0.003 * |
| **Anesthesia type** |  |  |  |
| Inhaled anesthetics, n (%) | 12/15 (80%) | 13/15 (87%) | — |
| TIVA, n (%) | 3/15 (20%) | 2/15 (13%) | — |

Baseline characteristics of the investigated cohort with myalgic encephalomyelitis/chronic fatigue syndrome (ME/CFS) as compared to the matched control cohort. BMI: body mass index; PONV: postoperative nausea and vomiting; ASA = American Society of Anesthesiologists; TIVA: total intravenous anesthesia. Continuous variables are presented as median and interquartile range (IQR); binary variables as n/N (%); — = not tested (matching variable or descriptive comparison only). \* $p < 0.05$ .

No previous anesthesia-related complications were documented in any of the included patients. A history of postoperative nausea and vomiting was present in 4 of 15 ME/CFS patients (27%) and 2/15 controls (13%). Preoperative regional anesthesia was performed in one patient in each group. Anesthesia and surgical duration, as well as time from end of surgery to extubation, were similar between groups. Postoperative hospital length also did not differ between groups.

### 3.2. Anesthetic management and intraoperative hemodynamics

Parameters of anesthetic management did not differ between groups (Table 2). Sufentanil was administered in 5 ME/CFS patients and 6 controls; Remifentanil was used in 10 ME/CFS patients and 9 controls. Two cases per group were excluded from remifentanil infusion rate comparisons (n=1 per group administered via target-controlled infusion, n=1 per group documented in µg·kg⁻¹·min⁻¹), leaving 13 analyzable pairs. Piritramide was used in 3 ME/CFS patients and no controls, while oxycodone was given in 6 ME/CFS patients (40%) and 8 controls (53%). Neither intraoperative piritramide nor oxycodone use differed significantly between groups. End-tidal volatile anesthetic concentrations (in minimum alveolar concentration (MAC) equivalents) were similar between groups. Intraoperative fluid administration and vasopressor management, including the proportion of patients receiving vasopressors and the maximum noradrenaline infusion rate, were comparable between groups.

**Table 2.**
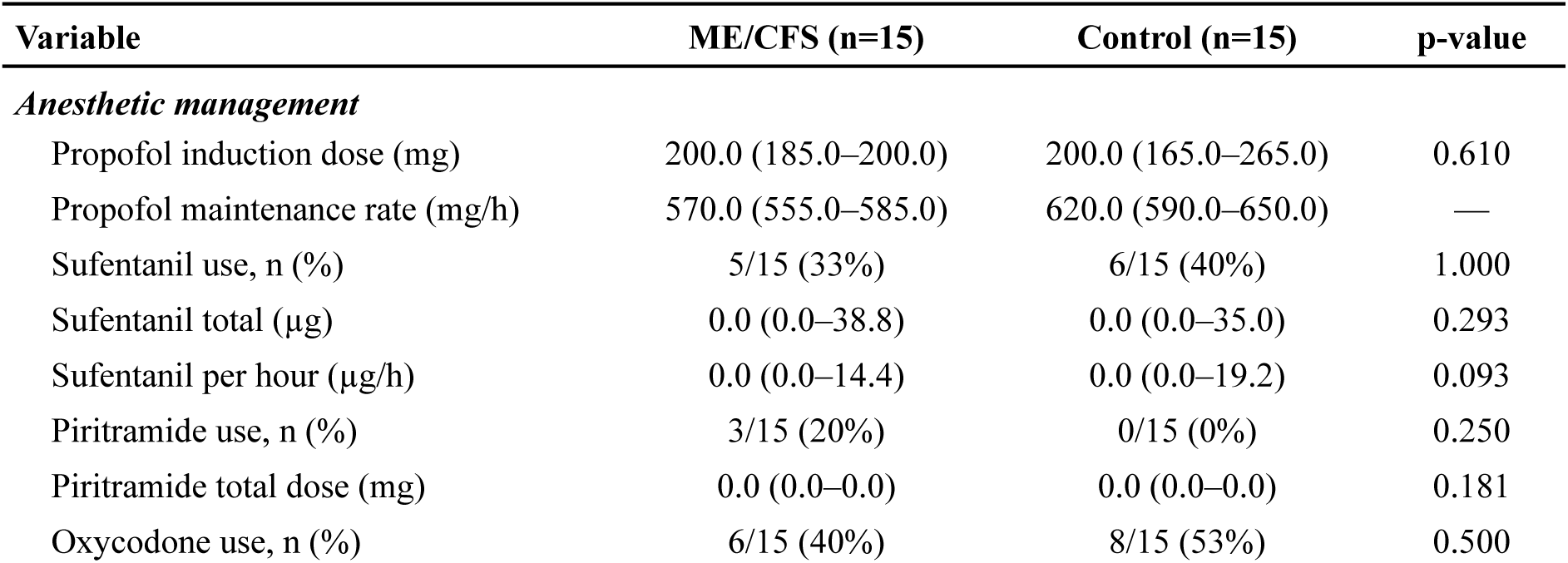
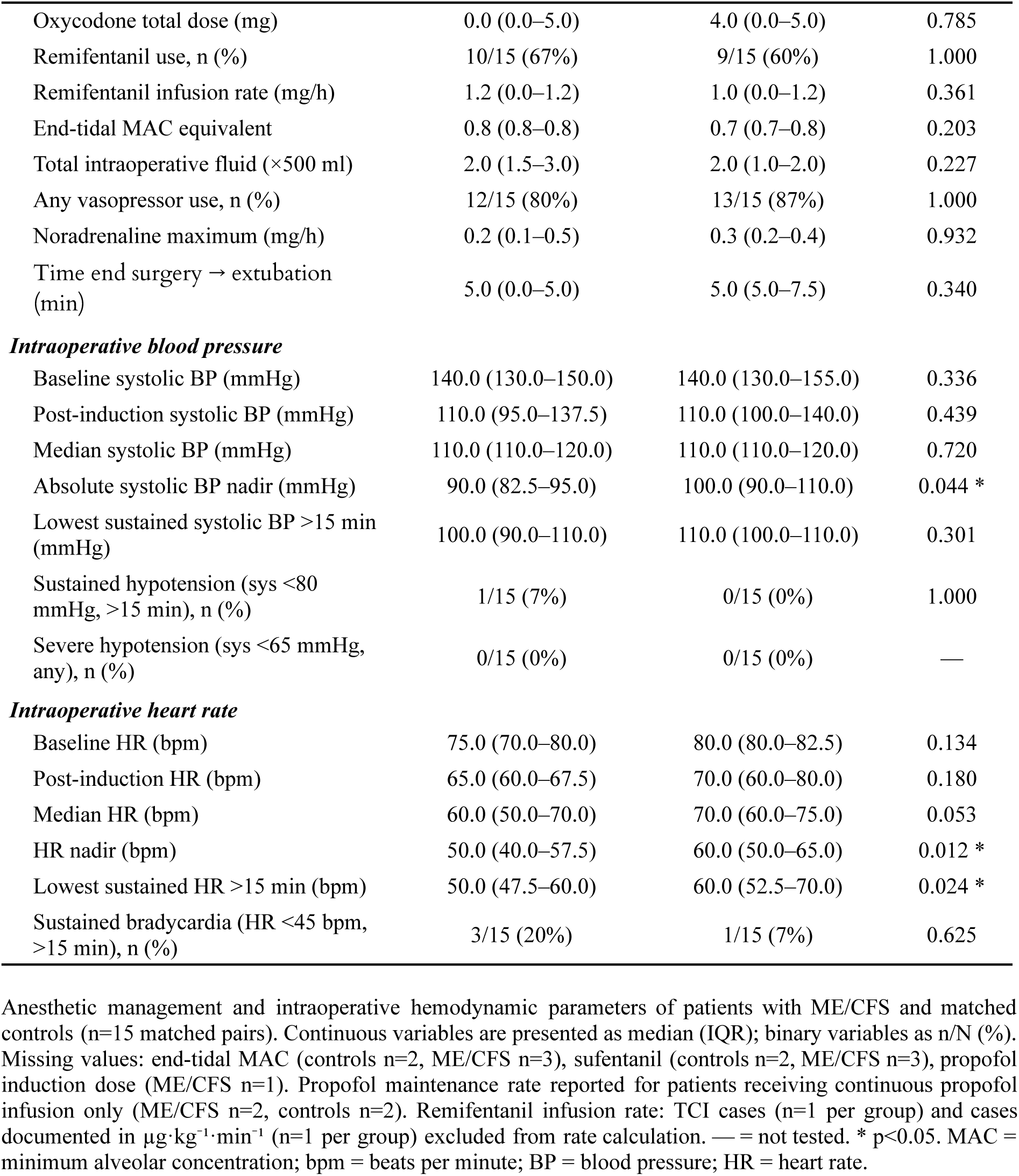
Anesthetic management and intraoperative hemodynamics.

**Table 3.** Post-anesthesia care unit outcomes.

| Variable | ME/CFS (n=15) | Control (n=15) | p-value |
| --- | --- | --- | --- |
| **Pain** |  |  |  |
| Maximum NRS pain score (0–10) | 5.0 (4.0–6.0) | 1.0 (0.0–4.0) | 0.008 * |
| **Analgesia** |  |  |  |
| Opioid rescue analgesia, n (%) | 12/15 (80%) | 5/15 (33%) | 0.039 * |
| Non-opioid rescue analgesia, n (%) | 6/15 (40%) | 5/15 (33%) | 1.000 |
| **Other complications** |  |  |  |
| PONV event, n (%) | 1/15 (7%) | 2/15 (13%) | 1.000 |
| Oxygen supplementation at discharge, n (%) | 7/15 (47%) | 5/15 (33%) | 0.625 |
| **Recovery** |  |  |  |
| PACU stay duration (min) | 60.0 (52.5–95.0) | 70.0 (57.5–85.0) | 0.861 |
| **Vital signs in the PACU** |  |  |  |
| Systolic BP at admission (mmHg) | 130.0<br>(120.0–160.0) | 130.0<br>(120.0–147.5) | 0.286 |
| Systolic BP at discharge (mmHg) | 130.0<br>(120.0–140.0) | 130.0<br>(120.0–140.0) | 1.000 |
| HR at admission (bpm) | 90.0 (81.2–90.0) | 80.0 (70.0–90.0) | 0.203 |
| HR at discharge (bpm) | 80.0 (80.0–82.5) | 70.0 (60.0–86.2) | 0.078 |

Post-anesthesia care unit (PACU) outcomes in patients with myalgic encephalomyelitis/chronic fatigue syndrome (ME/CFS) and matched controls (n=15 matched pairs). Continuous variables are presented as median (IQR); binary variables as n/N (%). Missing values: PACU vital signs at admission (controls n=1), heart rate at discharge (controls n=3, ME/CFS n=3). BP = blood pressure; bpm = beats per minute; HR = heart rate; IQR = interquartile range; mmHg = millimeters of mercury; NRS = numerical rating scale; PACU = post-anesthesia care unit; PONV = postoperative nausea and vomiting. \* p<0.05.

Intraoperative hemodynamic parameters were largely similar between groups (Table 2). Pre-induction blood pressure, post-induction blood pressure, and median intraoperative blood pressure did not differ significantly. However, the lowest recorded systolic blood pressure was significantly lower in ME/CFS patients compared with controls (median 90.0 [IQR 82.5-95.0] vs. 100.0 [IQR 90.0-110.0] mmHg; p=0.044; Fig. 2-A), as was the lowest recorded heart rate (median 50.0 [IQR 40.0-57.5] vs. 60.0 [IQR 50.0-65.0] bpm; p=0.012; Fig. 2-B). Importantly, no episodes of severe hypotension (systolic blood pressure <65 mmHg) occurred in either group.

**Fig. 2:**
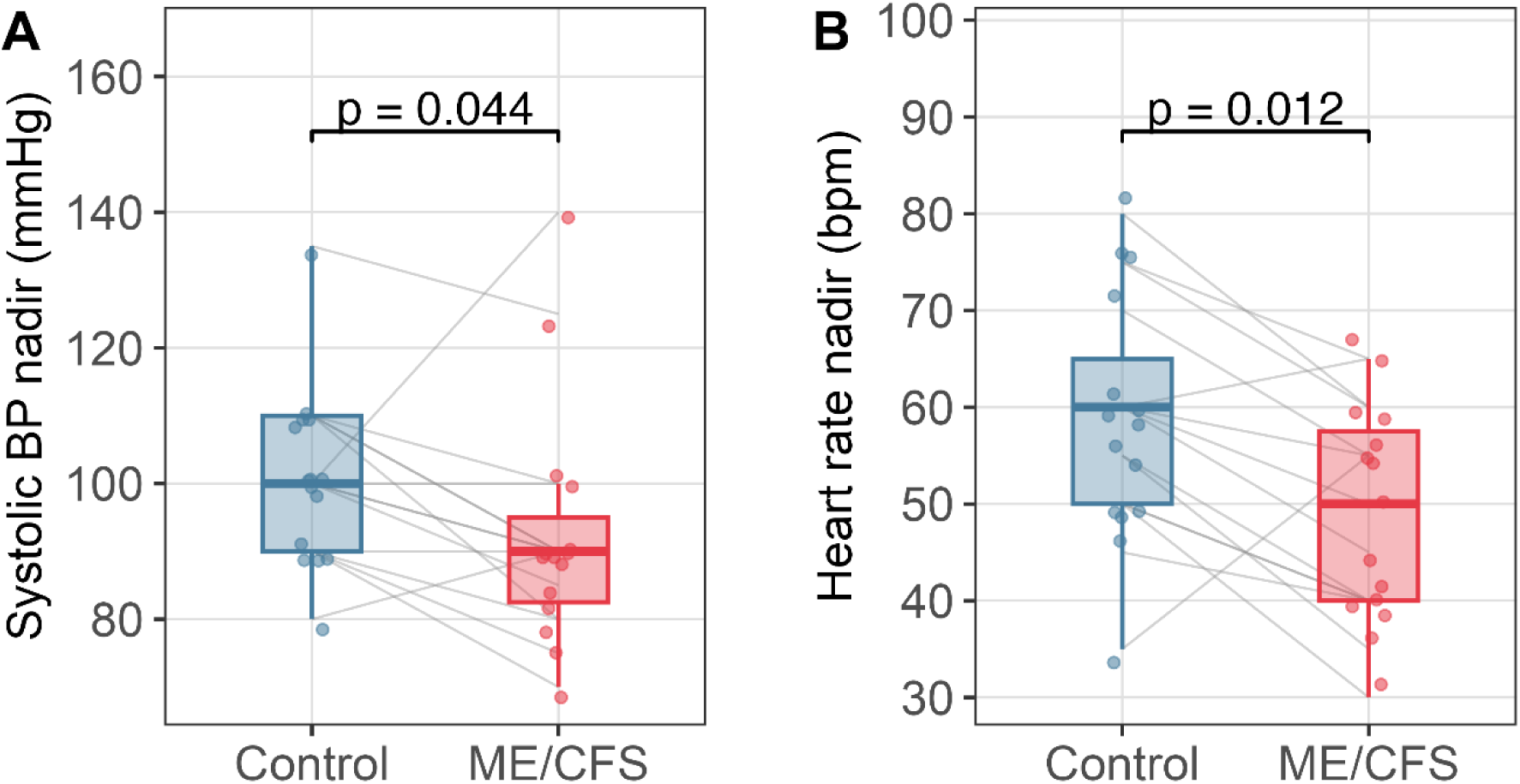
Boxplots of lowest recorded intraoperative systolic blood pressure (A) and heart rate (B) in patients with myalgic encephalomyelitis/chronic fatigue syndrome (ME/CFS) and matched controls undergoing general anesthesia (n=15 matched pairs). Lines connect matched pairs.

### 3.3. Post-anesthesia care unit outcomes

In the PACU, ME/CFS patients reported significantly higher maximum pain scores compared with controls (Fig. 3-A).

**Fig. 3:**
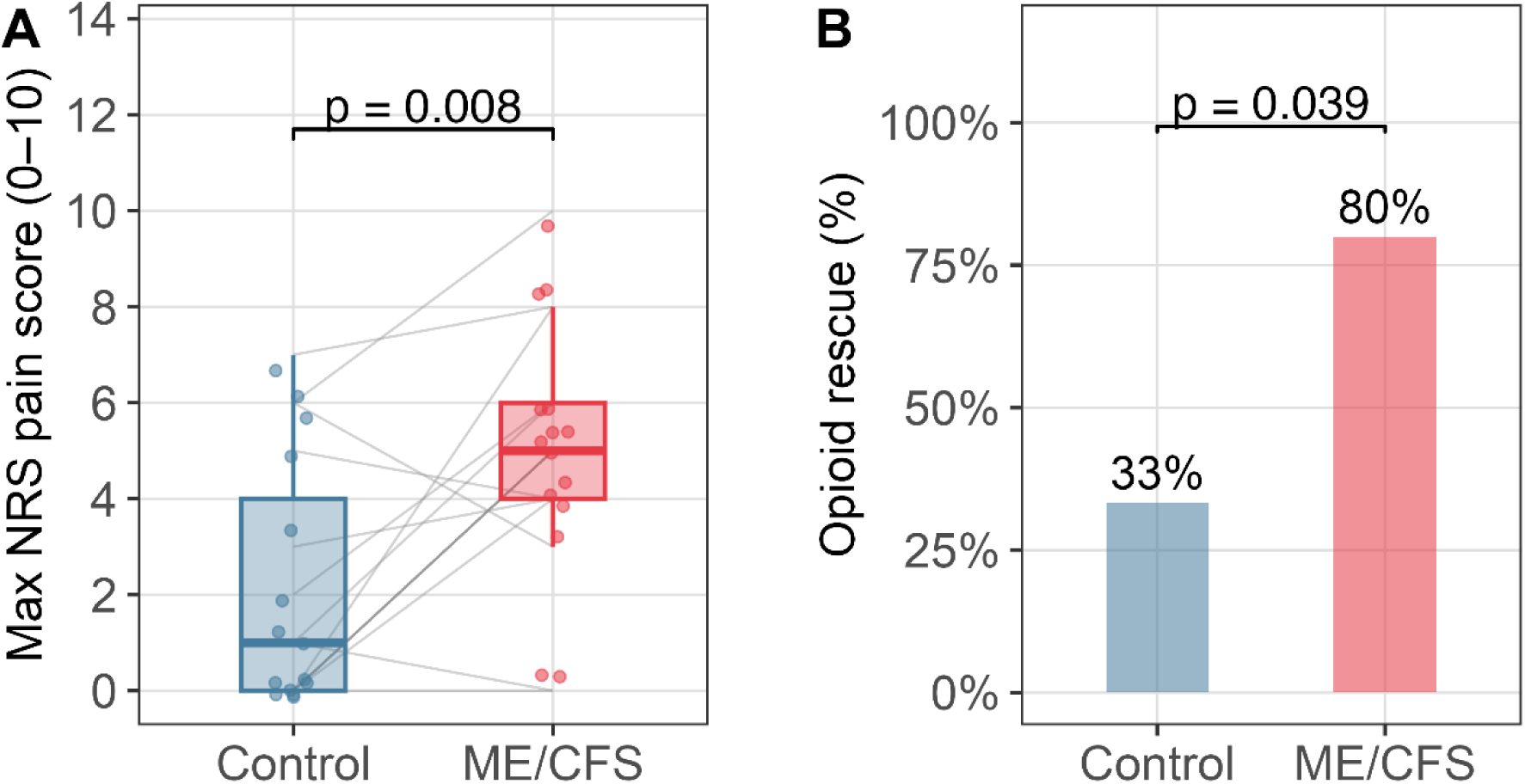
Post-anesthesia care unit pain ratings in patients with myalgic encephalomyelitis/chronic fatigue syndrome (ME/CFS) and matched controls after general anesthesia (n=15 matched pairs). **(A)** Maximum numerical rating scale (NRS) pain score recorded during post-anesthesia care (0 = no pain, 10 = worst imaginable pain). **(B)** Proportion of patients requiring opioid rescue analgesia. Lines connect matched pairs.

Opioid rescue analgesia was required in 12 of 15 ME/CFS patients (80%) compared with 5 of 15 control patients (33%), representing a statistically significant difference (p = 0.039), (Fig. 3-B). The absolute risk difference for opioid rescue analgesia was 47%.

In one ME/CFS patient undergoing lower-limb debridement, postoperative pain remained poorly controlled despite systemic analgesia and could only be adequately controlled after placement of a distal sciatic nerve catheter in the PACU.

Other PACU outcomes, including non-opioid rescue analgesia, oxygen supplementation, postoperative nausea and vomiting, and PACU length of stay, did not differ significantly between groups.

## 4. Discussion

This retrospective matched-pair analysis provides clinical data on perioperative outcomes in individuals with myalgic encephalomyelitis/chronic fatigue syndrome (ME/CFS) undergoing general anesthesia. ME/CFS represents a potentially vulnerable population that remains underrepresented in the anesthesia literature and for whom evidence-based perioperative guidance is currently lacking. Overall, general anesthesia appeared to be well tolerated from a hemodynamic perspective, whereas postoperative recovery, particularly pain, was more adversely affected compared with matched controls.

Intraoperatively, individuals with ME/CFS exhibited lower lowest intraoperative systolic blood pressure and heart rate values compared with matched controls. However, these differences did not translate into clinically relevant hemodynamic instability. No severe adverse events or escalation of therapy were observed. Vasopressor use, fluid administration, and other hemodynamic parameters were comparable between groups, and no episodes of clinically significant hypotension occurred. Continuous low-dose noradrenaline infusions are routinely used in our institution during general anesthesia, including in low-risk patients, which may have contributed to the overall hemodynamic stability observed. In addition, no prolonged emergence from anesthesia was noted. However, given the small sample size, the study is not powered to reliably assess the incidence of rare perioperative adverse events, and these findings should therefore be interpreted as hypothesis-generating rather than confirmatory.

In contrast, postoperative recovery appeared more affected. Individuals with ME/CFS reported higher pain scores and required opioid rescue analgesia more frequently in the PACU. In one case, analgesic therapy had to be escalated with placement of a distal sciatic nerve catheter due to insufficient pain control with systemic medication alone. These findings indicate a clinically relevant difference in early postoperative pain and may have implications for perioperative analgesic strategies in this population.

The observed hemodynamic differences may reflect underlying autonomic dysfunction, a common feature of ME/CFS. A subset of patients presents with postural orthostatic tachycardia syndrome, which may predispose to altered cardiovascular responses under anesthesia, including exaggerated vagal responses or impaired sympathetic compensation. Consistent with this, intraoperative hypotension has been reported in patients with POTS undergoing general anesthesia [7]. However, in our cohort, these physiological differences did not translate into clinically relevant instability. In contrast, the increased postoperative pain burden may be related to altered nociceptive processing. Central sensitization has been documented in a substantial proportion of individuals with ME/CFS and may represent a plausible mechanism underlying the observed differences [15]. Alternatively, pre-existing chronic pain or other unmeasured factors may have contributed, as preoperative pain scores were not available.

Overall, evidence on anesthesia in ME/CFS remains extremely limited. Our findings are broadly consistent with the existing literature [27], suggesting that standard anesthetic approaches may be hemodynamically well tolerated, although robust data remain lacking. Importantly, patient-relevant outcomes such as post-exertional malaise and symptom exacerbation remain largely unstudied and require dedicated investigation. The potential impact of anesthetic techniques and perioperative management strategies in ME/CFS remains largely unexplored. Data on alternatives to general anesthesia, such as regional or neuraxial techniques alone or in combination with sedation or general anesthesia, are lacking. It is conceivable that the choice of anesthetic technique, pharmacological regimen, and perioperative care strategies may influence not only immediate postoperative outcomes but also medium- and longer-term symptom trajectories, including the risk of symptom exacerbation.

Crucially, post-exertional malaise - the hallmark feature of ME/CFS [28] - typically manifests with delayed onset and is not captured by routine perioperative documentation focused on immediate outcomes. This has important implications not only for the assessment of anesthetic techniques and perioperative management strategies, but also for clinical practice, particularly in the context of repeated or staged interventions, where patients may undergo subsequent procedures while already in a state of post-exertional malaise. The increased postoperative pain and need for opioid analgesics in the ME/CFS cohort might be part of PEM. Prospective studies incorporating longitudinal follow-up are therefore essential to adequately capture this dimension of perioperative risk.

This study has several limitations. The retrospective single-center design, reliance on handwritten anesthetic records with five-minute hemodynamic documentation, and small sample size of 15 matched pairs limit statistical power and constrain the conclusions that can be drawn, particularly regarding rare adverse events. Case identification based on ICD-10 codes without systematic verification against established diagnostic criteria such as the Canadian Consensus Criteria [22] introduces potential misclassification. In addition, the number of identified cases was substantially lower than expected based on population prevalence estimates, likely reflecting both underdiagnosis of ME/CFS and limitations of coding-based case identification, which may fail to capture diagnoses documented in clinical correspondence but not formally coded. Patients with ME/CFS may also avoid elective procedures due to concerns about post-exertional malaise, further contributing to underrepresentation. Consequently, the true perioperative burden of ME/CFS is likely underestimated. However, the objective of this study was not to provide precise incidence estimates or fully characterize the perioperative ME/CFS population - which would require substantially larger, systematically ascertained cohorts - but to identify perioperative patterns and signals warranting further investigation. These limitations therefore primarily affect generalizability, while internal comparison remains informative. Selection bias is inherent to surgical cohorts, as more severely affected individuals are less likely to undergo elective procedures. Preoperative pain scores were not available, precluding adjustment for baseline differences, and post-exertional malaise - arguably the most clinically relevant outcome in ME/CFS - was not assessed. In addition, the study was not powered to detect small-to-moderate effect sizes, and non-significant findings should not be interpreted as evidence of equivalence. Finally, findings from this single-center cohort may not be generalizable to other settings or to ME/CFS populations with different severity profiles or comorbidity patterns.

Despite these limitations, this exploratory matched analysis provides clinically relevant signals in a field with very limited existing evidence and identifies key areas for future prospective studies aimed at improving perioperative management in individuals with ME/CFS.

## 5. Conclusion

This retrospective matched analysis provides clinical data on perioperative responses in individuals with ME/CFS, a population for whom evidence in anesthesia is extremely limited. General anesthesia appeared hemodynamically well tolerated, although lower lowest intraoperative hemodynamic values suggest altered physiological responses without clear clinical consequences. In contrast, postoperative pain burden substantially increased, indicating a potential vulnerability requiring careful analgesic strategies. Future studies should address the influence of anesthetic techniques and evaluate post-exertional malaise as a key disease feature, including its relevance for perioperative management and delayed outcomes. Our findings support the need for prospective studies to better characterize perioperative risk and optimize anesthetic care in this population.

## Ethics

This study was approved by the Ethics Committee of the University of Regensburg (reference number: 25-4271-101). The requirement for informed consent was waived in accordance with institutional and national regulations due to the retrospective design and use of routinely collected, de-identified clinical data.

## CRediT authorship contribution statement

**Felix Steinkirchner:** Conceptualization, Data curation, Formal analysis, Investigation, Methodology, Project administration, Visualization, Writing – original draft, Writing – review & editing. **Christina Kaufmann:** Conceptualization, Data curation, Investigation, Writing – review & editing. **Richard Felix Kraus:** Investigation, Writing – review & editing. **Maximilian Käss:** Investigation, Writing – review & editing. **Elisabeth Schieffer:** Conceptualization, Supervision, Writing – review & editing. **Bernhard M. Graf:** Resources, Writing – review & editing. **Christoph Lassen:** Conceptualization, Supervision, Writing – review & editing. **Viktoria Kimmerling:** Conceptualization, Investigation, Writing – review & editing. **Alexander Dejaco:** Conceptualization, Formal analysis, Funding acquisition, Investigation, Supervision, Writing – original draft, Writing – review & editing.

## Declaration of generative AI use

During the preparation of this work, the authors used ChatGPT (OpenAI) and Claude (Anthropic) to assist with language editing and to improve readability. The authors reviewed and edited all outputs and take full responsibility for the content of this manuscript.

## Data statement

The data that support the findings of this study are not publicly available due to data protection and privacy regulations but are available from the corresponding author upon reasonable request, subject to applicable data protection regulations.

## Data Availability

The data that support the findings of this study are available from the authors upon reasonable request; however, only data that can be shared in accordance with applicable data protection and privacy regulations will be provided.

