## Supplemental Material for "Perioperative outcomes in patients with myalgic encephalomyelitis/chronic fatigue syndrome undergoing general anesthesia: a retrospective matched-pair study"

* Corresponding author

### Supplementary material

**Supplementary Table S1. Characteristics of matched pairs**

| Diagnosis | Procedure | Sex | ASA | Year |
|---|---|---|---|---|
| **ME/CFS** | **Ophthalmic surgery (phacoemulsification, IOL implantation, ppV)** | **F** | **II** | **2015** |
| Control | Ophthalmic surgery (phacoemulsification, IOL implantation, ppV) | F | II | 2015 |
| **ME/CFS** | **Pansinusotomy** | **F** | **II** | **2016** |
| Control | Pansinusotomy | F | II | 2018 |
| **ME/CFS** | **Dental extraction** | **F** | **III** | **2017** |
| Control | Mandibular cystectomy | F | II | 2018 |
| **ME/CFS** | **Hepatic resection** | **F** | **III** | **2018** |
| Control | Hepatic resection | F | II | 2018 |
| **ME/CFS** | **Hemithyroidectomy** | **F** | **II** | **2022** |
| Control | Hemithyroidectomy | F | II | 2025 |
| **ME/CFS** | **Total hip arthroplasty** | **F** | **II** | **2023** |
| Control | Total hip arthroplasty | F | II | 2025 |
| **ME/CFS** | **Tympanoplasty** | **M** | **II** | **2023** |
| Control | Tympanoplasty | M | I | 2025 |
| **ME/CFS** | **Pterygomandibular abscess drainage** | **M** | **II** | **2024** |
| Control | Pterygomandibular abscess drainage | M | II | 2025 |
| **ME/CFS** | **Ventriculoperitoneal shunt placement** | **F** | **III** | **2024** |
| Control | Ventriculoperitoneal shunt placement | F | III | 2025 |
| **ME/CFS** | **MRI brain under general anesthesia** | **F** | **III** | **2024** |
| Control | MRI brain under general anesthesia | F | II | 2025 |
| **ME/CFS** | **Pansinusotomy** | **M** | **II** | **2024** |
| Control | Pansinusotomy | M | III | 2025 |
| **ME/CFS** | **VATS** | **F** | **III** | **2024** |
| Control | VATS | F | III | 2025 |
| **ME/CFS** | **Lower limb soft tissue debridement** | **F** | **III** | **2025** |
| Control | Lower limb soft tissue debridement | F | III | 2025 |
| **ME/CFS** | **Laparoscopic cholecystectomy** | **F** | **II** | **2025** |

| Diagnosis | Procedure | Sex | ASA | Year |
|---|---|---|---|---|
| Control | Laparoscopic cholecystectomy | F | II | 2026 |
| **ME/CFS** | **Ophthalmic surgery (ppV, membrane peeling)** | **M** | **III** | **2026** |
| Control | Ophthalmic surgery (phacoemulsification, IOL implantation) | M | III | 2026 |

Baseline and procedural characteristics of all 15 matched pairs. Each pair consists of one patient with myalgic encephalomyelitis/chronic fatigue syndrome (ME/CFS, bold) and one matched control. Pairs are ordered by year of the ME/CFS procedure. Matching was performed on sex and procedure type; age, BMI, and year of procedure were additionally considered to avoid large discrepancies. ASA: American Society of Anesthesiologists physical status classification; BMI: body mass index; IOL: intraocular lens implantation; MRI: magnetic resonance imaging; VATS: video-assisted thoracoscopic surgery; ppV: pars plana vitrectomy.

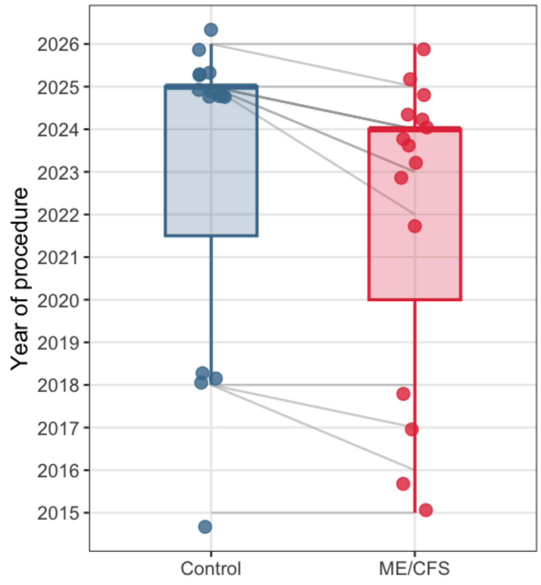

**Figure S1.** Year of procedure for myalgic encephalomyelitis/chronic fatigue syndrome (ME/CFS) patients and matched controls (n=15 matched pairs). Each box represents the median and interquartile range; individual data points are shown as dots; lines connect matched pairs. ME/CFS: myalgic encephalomyelitis/chronic fatigue syndrome.
